# Attributional bias in epilepsy: differences between genetic generalised epilepsy, temporal lobe epilepsy and healthy controls

**DOI:** 10.64898/2026.04.28.26351955

**Authors:** Viktoria Pytelova, Ela Gatialova, Jakub Zalud, Martin Modrak, Eliska Ksirova, Michaela Kalinova, Adam Kalina, Petr Marusic, Jana Amlerova

## Abstract

**Background:** Attributional bias, a tendency to overinterpret others’ intentions as hostile (rather than situational or accidental), represents a component of social cognition and may affect everyday functioning. Neural models link attributional processing to fronto-temporal circuits and the default mode network, which are frequently altered in epilepsy. Difficulties in social participation and employment are common in people with epilepsy, and maladaptive attributional styles may contribute to these challenges. Attributional bias has not been systematically compared across epilepsy syndromes.

**Methods:** We examined attributional bias in 96 participants comprising 26 individuals with genetic generalised epilepsy (GGE), 27 with temporal lobe epilepsy (TLE), and 43 healthy controls (HC). Attributional style was assessed using the Ambiguous Intentions Hostility Questionnaire. Depressive symptoms were evaluated using the Neurological Disorders Depression Inventory in Epilepsy. Group differences were analysed, and potential clinical and demographical correlates were explored.

**Results:** The GGE group exhibited higher hostility bias scores than HC (95% CI: 0.12–0.38, adjusted p = 0.014), whereas the difference between TLE and HC groups was moderate and not statistically significant (95% CI: 0.12–0.58, adjusted p = 0.059). Higher blame scores were positively associated with depressive symptoms (p = 0.016). Disease duration, seizure frequency, and antiseizure medication were not significantly associated with attributional bias.

**Conclusions:** These findings suggest that some individuals with genetic generalised epilepsy are more likely to interpret ambiguous situations as hostile. Altered attributional style may represent an under-recognised factor contributing to social difficulties in people with epilepsy and warrants further investigation as a potential target for psychosocial interventions.

**Highlights:**

- Some people with epilepsy are more prone to interpret social situations as hostile.
- Higher depression scores correlate with a tendency to blame external factors for misfortunes.
- Disease duration, antiseizure medication, and seizure frequency do not seem to influence the attributional bias.

## 1. INTRODUCTION

Attribution theory is a framework that explains how individuals interpret the causes of their own and others’ behaviours, attributing them to either internal dispositions or external circumstances. Internal (dispositional) attributions are based on personal characteristics, traits, or abilities, whereas external (situational) attributions are explanations based on external factors or the environment (Kestemont et al., 2015). Attribution theory also explores whether people attribute behaviours to stable or unstable causes (e.g., talent vs luck) and whether they view those causes as controllable or uncontrollable (e.g., effort vs genetic ability). Various cognitive biases can distort these attributions (Decombe et al., 2022). Attributional biases examine how one responds in certain social situations and identify extreme response patterns. Among these is hostility bias, a tendency to interpret others’ actions as intentional and hostile rather than accidental or due to chance. This misinterpretation of others’ motives can increase anger and hostility in the individual (Buck et al., 2017), thus leading to social maladaptation.

Studies on social competence in people with epilepsy (PWE) confirmed diminished self-esteem, strained interpersonal relationships, higher risk of emotional and behavioural problems, unemployment, and marital failure (Schachter, 2006; Yogarajah and Mula, 2019). The causes of social maladaptation in PWE are not fully understood and are likely multifactorial. Researchers approach the explanation of social maladaptation in PWE from various perspectives. Psychosocial factors, including epilepsy-related stigma or parental overprotectiveness, represent one point of view (Steiger and Jokeit, 2017; Yogarajah and Mula, 2019). Another angle arises from disease-related factors, including the duration of epilepsy, frequency of seizures, and side effects of anti-seizure medication (Hao et al., 2015; Zhang et al., 2018). Different standpoints emerge from psychiatric conditions of PWE, mainly depression, suicidal ideation, and anxiety that occur with higher frequency in PWE than in the general population and could also promote social ineptitude (Kanner, 2022). Social cognition deficits offer yet another distinct perspective on the reduced social skills in PWE.

Social cognition, one of the six core neurocognitive domains (American Psychiatric Association, 2013), encompasses the processes involved in perceiving, interpreting, and responding to social information. It refers to how people think about themselves and others in social contexts (Fiske and Taylor, 2013). This construct has several dimensions, including the Theory of Mind (ToM), emotion recognition (ER), social perception, social knowledge, and attributional bias (Decombe et al., 2022). Neurobiologically, the social brain network involves dynamic collaboration among regions in the frontal and temporal lobes, with differences in structural involvement depending on the measurement methodology (data-driven or task-driven)(Krendl and Betzel, 2022). Advanced neuroimaging studies suggest that the default mode network (DMN) plays a central role in social cognitive processes (Krendl and Betzel, 2022). Inferring about the motivations of other people and attributing causes to their behaviour activates brain areas that highly overlap with the DMN. (Kestemont et al., 2015) Altered connectivity in the DMN has been described in both TLE and IGE (Parsons et al., 2020; Zanão et al., 2021).

Previous research on social cognition impairment in PWE has focused primarily on focal syndromes, like frontal lobe epilepsy (Farrant et al., 2005; Ziaei et al., 2022) and predominantly temporal lobe epilepsy (TLE) (Amlerova et al., 2014; Broicher et al., 2012; Giovagnoli et al., 2021; Jokeit, 2010; Mirabel et al., 2020). Frequently studied concepts include ER and ToM. Individuals with TLE exhibit moderate-to-severe deficits in ER and ToM (Ziaei et al., 2022). As we noted in our recent review (Ogurcakova et al., 2024), evidence for social cognition deficits in individuals with generalised epilepsy is scarce and inconsistent, possibly due to the vast phenotypical heterogeneity within the generalised epilepsy group and even within a single syndrome, e.g., juvenile myoclonic epilepsy. Some studies suggest a potential deficit in social skills among children with idiopathic generalised epilepsy (IGE) (Berg et al., 2007; Stewart et al., 2019). However, these deficits did not correlate with facial ER impairments or epilepsy-related variables (age of seizure onset, duration of epilepsy, seizure frequency, epilepsy severity, or number of antiseizure drugs). Studies on attributional style among epilepsy patients are scarce, and either do not distinguish among epilepsy syndromes (Gehlert, 1994) or focus on TLE only (Hermann et al., 1996).

This study aimed to investigate attributional biases in genetic generalised epilepsy (GGE) and TLE populations and compare them with those of a healthy control group. We hypothesised that attributional biases would be more pronounced in both TLE and GGE groups than in HC, with higher scores in the TLE group and greater heterogeneity in GGE, consistent with previously reported Theory of Mind impairments.

## 2. METHODS

### Healthy controls

We created the healthy control (HC) group by combining volunteer hospital employees, medical students, and individuals who responded to an online advertisement seeking research assistance at our facility. Participants were not offered any reward, and all signed an informed consent form approved by the local ethics committee. We excluded volunteers who had been treated for neurologic or psychiatric disorders.

### People with epilepsy

We asked subjects who fulfilled the criteria to participate in the study during routine clinic visits. All participants gave informed consent.

Inclusion criteria:

- Electro-clinical diagnosis of either GGE or TLE

Exclusion criteria:

- Epilepsy surgery in history, including vagal nerve stimulation or deep brain stimulation (these interventions could influence the cognitive profile of PWE and could introduce bias into attributional styles testing results)
- History of psychosis or schizophrenia (both diagnoses involve a risk for hostile attributional bias)
- Severe cognitive deficit (e.g., neurodevelopmental cognitive disability preventing the patient from understanding test instructions)

We examined the participants between March 2023 and November 2023. The data collection period was constrained by a specific time limit set by a grant support deadline (JUNIOR grant; see acknowledgements). We also collected sociodemographic data (age, sex, and education) for descriptive purposes. To explore possible confounders, we collected epilepsy-related variables, including disease duration, antiseizure medication use, and seizure occurrence in the 30 days preceding the examination. Since depressive symptoms are known to influence attributional style and possibly exaggerate hostility (Smith et al., 2016), we measured depressive symptoms using the Czech version of the Neurological Disorders Depression Inventory for Epilepsy (NDDI-E). See Table 1a and Table 1b. For further, more detailed clinical characteristics of the GGE and TLE groups, see the supplementary data.

**Table 1a.**
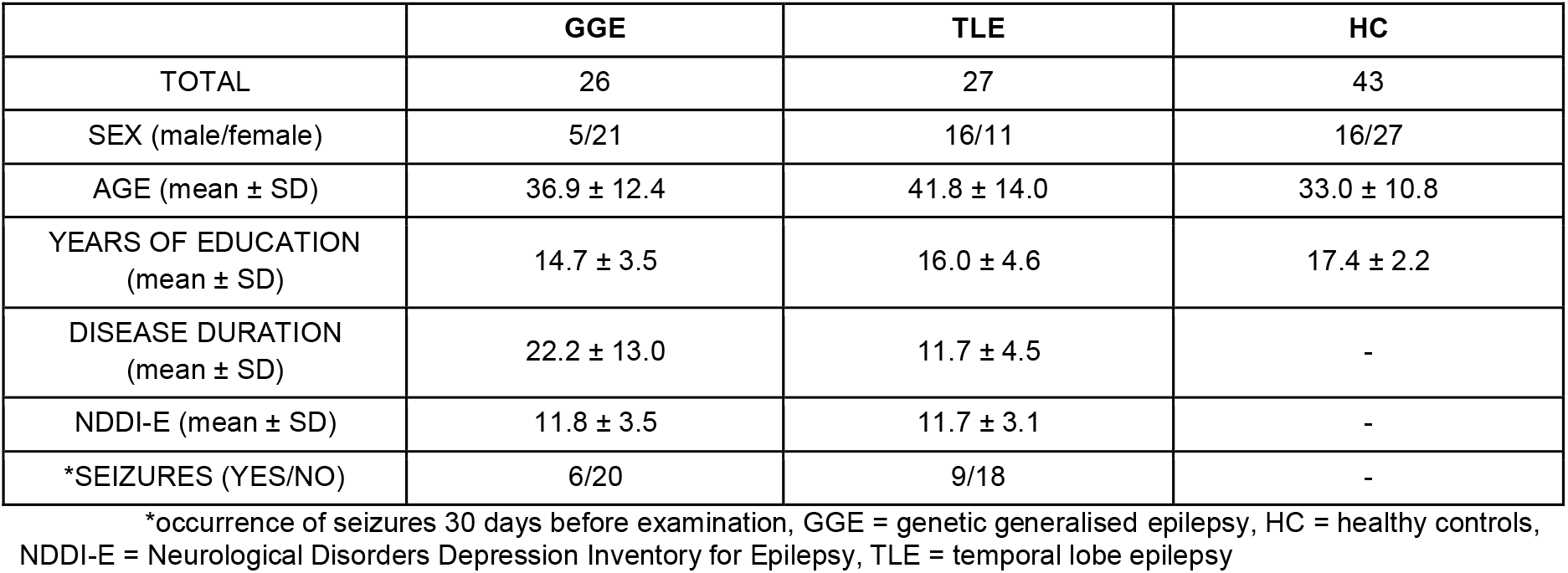

**Table 1b.**
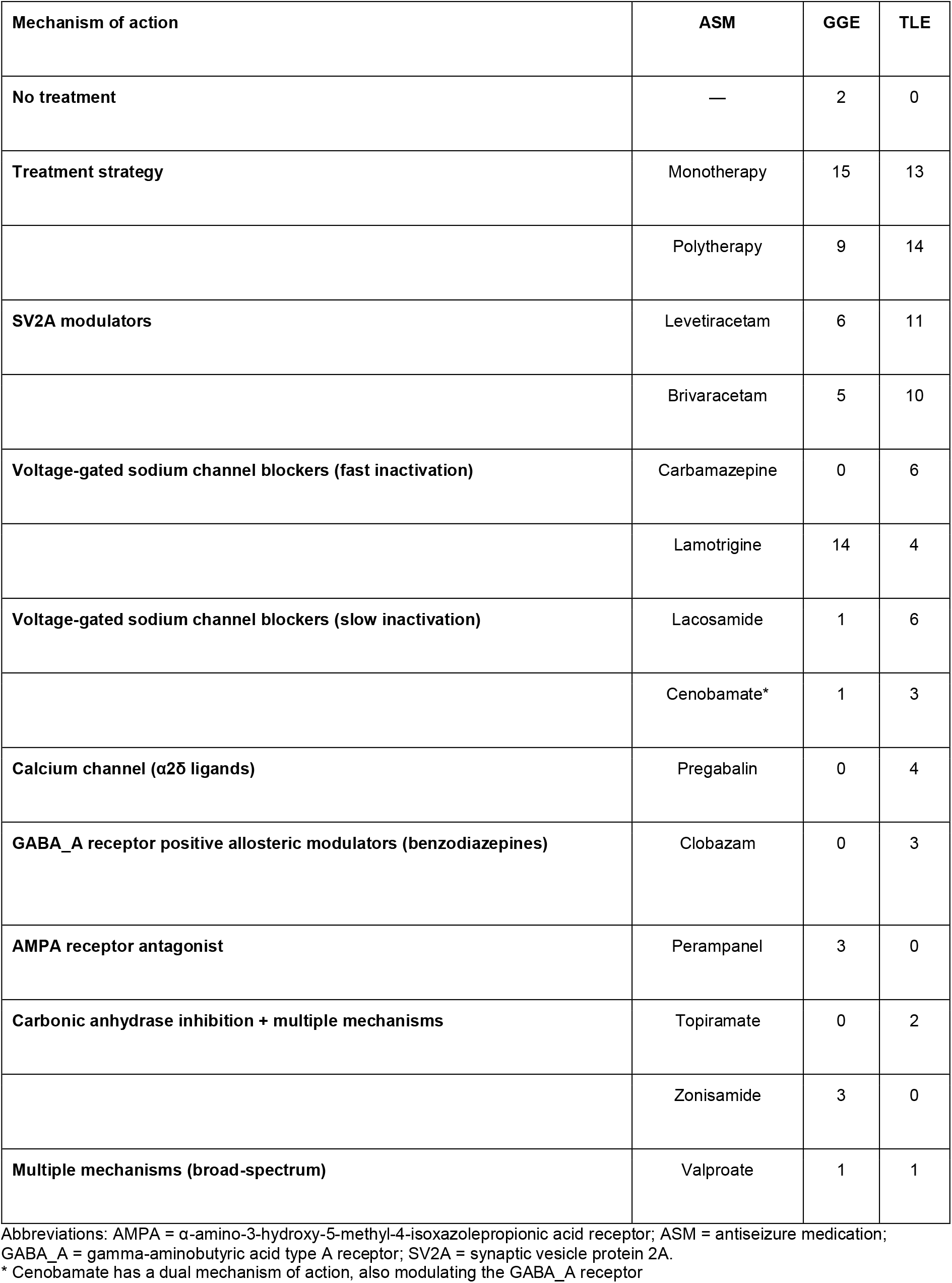

| Mechanism of action | ASM | GGE | TLE |
| --- | --- | --- | --- |
| No treatment | — | 2 | 0 |
| Treatment strategy | Monotherapy | 15 | 13 |
|  | Polytherapy | 9 | 14 |
| SV2A modulators | Levetiracetam | 6 | 11 |
|  | Brivaracetam | 5 | 10 |
| Voltage-gated sodium channel blockers (fast inactivation) | Carbamazepine | 0 | 6 |
|  | Lamotrigine | 14 | 4 |
| Voltage-gated sodium channel blockers (slow inactivation) | Lacosamide | 1 | 6 |
|  | Cenobamate* | 1 | 3 |
| Calcium channel ($\alpha 2\delta$ ligands) | Pregabalin | 0 | 4 |
| GABA<sub>A</sub> receptor positive allosteric modulators (benzodiazepines) | Clobazam | 0 | 3 |
| AMPA receptor antagonist | Perampanel | 3 | 0 |
| Carbonic anhydrase inhibition + multiple mechanisms | Topiramate | 0 | 2 |
|  | Zonisamide | 3 | 0 |
| Multiple mechanisms (broad-spectrum) | Valproate | 1 | 1 |

Abbreviations: AMPA = $\alpha$ -amino-3-hydroxy-5-methyl-4-isoxazolepropionic acid receptor; ASM = antiseizure medication; GABA<sub>A</sub> = gamma-aminobutyric acid type A receptor; SV2A = synaptic vesicle protein 2A.
\* Cenobamate has a dual mechanism of action, also modulating the GABA<sub>A</sub> receptor

**Table 2:** Results overview.

|  | GGE (mean ± SD) | TLE (mean ± SD) | HC (mean ± SD) |
| --- | --- | --- | --- |
| **Hostility bias** |  |  |  |
| Total | 1.63 ± 0.59 | 1.56 ± 0.53 | 1.38 ± 0.47 |
| Ambiguous situations | 1.69 ± 0.50 | 1.61 ± 0.48 | 1.34 ± 0.38 |
| Intentional situations | 2.11 ± 0.48 | 1.97 ± 0.48 | 1.77 ± 0.47 |
| Accidental situations | 1.08 ± 0.13 | 1.10 ± 0.15 | 1.03 ± 0.09 |
| **Aggression bias** |  |  |  |
| Total | 1.81 ± 0.56 | 1.77 ± 0.53 | 1.67 ± 0.55 |
| Ambiguous situations | 1.65 ± 0.43 | 1.59 ± 0.42 | 1.54 ± 0.33 |
| Intentional situations | 2.28 ± 0.54 | 2.08 ± 0.46 | 2.04 ± 0.55 |
| Accidental situations | 1.49 ± 0.38 | 1.63 ± 0.58 | 1.58 ± 0.50 |
| **Blame score** |  |  |  |
| Total | 2.73 ± 0.89 | 2.64 ± 0.87 | 2.73 ± 0.83 |
| Ambiguous | 2.58 ± 0.62 | 2.43 ± 0.58 | 2.47 ± 0.47 |
| Intentional situations | 3.58 ± 0.81 | 3.47 ± 0.79 | 3.63 ± 0.62 |
| Accidental situations | 2.04 ± 0.39 | 2.06 ± 0.46 | 2.10 ± 0.44 |

GGE = genetic generalised epilepsy, TLE = temporal lobe epilepsy, HC = healthy controls,

**Table 3a:**
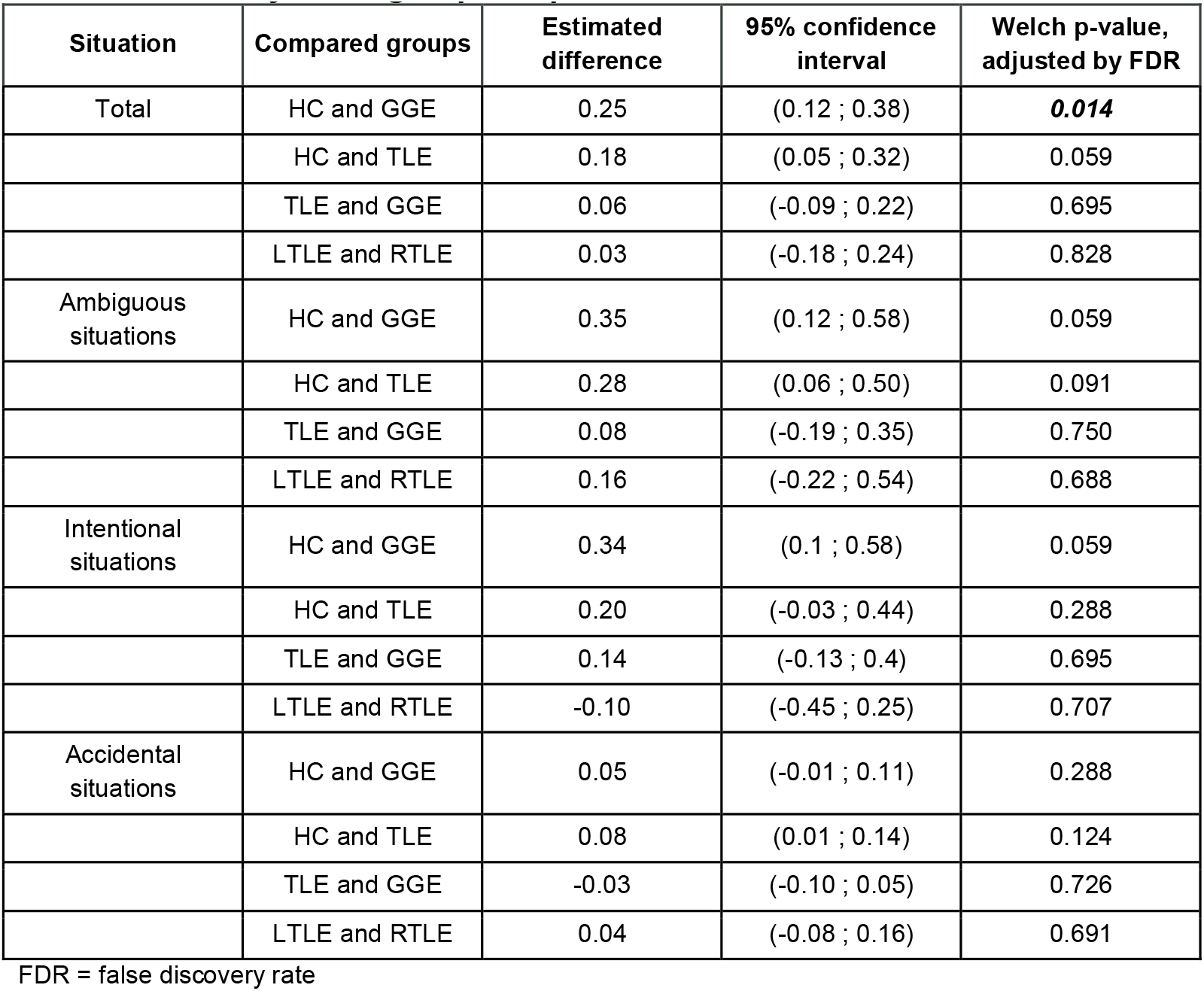
Hostility bias - group comparison.

| Situation | Compared groups | Estimated difference | 95% confidence interval | Welch p-value, adjusted by FDR |
| --- | --- | --- | --- | --- |
| Total | HC and GGE | 0.25 | (0.12 ; 0.38) | **0.014** |
|  | HC and TLE | 0.18 | (0.05 ; 0.32) | 0.059 |
|  | TLE and GGE | 0.06 | (-0.09 ; 0.22) | 0.695 |
|  | LTLE and RTLE | 0.03 | (-0.18 ; 0.24) | 0.828 |
| Ambiguous situations | HC and GGE | 0.35 | (0.12 ; 0.58) | 0.059 |
|  | HC and TLE | 0.28 | (0.06 ; 0.50) | 0.091 |
|  | TLE and GGE | 0.08 | (-0.19 ; 0.35) | 0.750 |
|  | LTLE and RTLE | 0.16 | (-0.22 ; 0.54) | 0.688 |
| Intentional situations | HC and GGE | 0.34 | (0.1 ; 0.58) | 0.059 |
|  | HC and TLE | 0.20 | (-0.03 ; 0.44) | 0.288 |
|  | TLE and GGE | 0.14 | (-0.13 ; 0.4) | 0.695 |
|  | LTLE and RTLE | -0.10 | (-0.45 ; 0.25) | 0.707 |
| Accidental situations | HC and GGE | 0.05 | (-0.01 ; 0.11) | 0.288 |
|  | HC and TLE | 0.08 | (0.01 ; 0.14) | 0.124 |
|  | TLE and GGE | -0.03 | (-0.10 ; 0.05) | 0.726 |
|  | LTLE and RTLE | 0.04 | (-0.08 ; 0.16) | 0.691 |

FDR = false discovery rate

**Table 3b:** Aggression bias - group comparison.

| Situation | Compared groups | Estimated difference | 95% confidence interval | Welch p-value, adjusted by FDR |
| --- | --- | --- | --- | --- |
| Total | HC and GGE | 0.14 | (-0.02 ; 0.29) | 0.288 |
|  | HC and TLE | 0.10 | (-0.08 ; 0.27) | 0.681 |
|  | TLE and GGE | 0.04 | (-0.15 ; 0.23) | 0.805 |
|  | LTLE and RTLE | 0.23 | (-0.06 ; 0.53) | 0.345 |
| Ambiguous situations | HC and GGE | 0.26 | (0.07 ; 0.46) | 0.072 |
|  | HC and TLE | 0.20 | (0.01 ; 0.40) | 0.176 |
|  | TLE and GGE | 0.06 | (-0.17 ; 0.30) | 0.750 |
|  | LTLE and RTLE | 0.12 | (-0.22 ; 0.46) | 0.691 |
| Intentional situations | HC and GGE | 0.24 | (-0.04 ; 0.51) | 0.288 |
|  | HC and TLE | 0.04 | (-0.20 ; 0.28) | 0.841 |
|  | TLE and GGE | 0.20 | (-0.08 ; 0.47) | 0.446 |
|  | LTLE and RTLE | 0.14 | (-0.22 ; 0.50) | 0.688 |
| Accidental situations | HC and GGE | -0.08 | (-0.30 ; 0.13) | 0.695 |
|  | HC and TLE | 0.05 | (-0.22 ; 0.33) | 0.813 |
|  | TLE and GGE | -0.14 | (-0.41 ; 0.14) | 0.695 |
|  | LTLE and RTLE | 0.44 | (-0.06 ; 0.94) | 0.307 |

FDR = false discovery rate

**Table 3c:** Blame score - group comparison.

| Situation | Compared groups | Estimated difference | 95% confidence interval | Welch p-value, adjusted by FDR |
| --- | --- | --- | --- | --- |
| Total | HC and GGE | 0.00 | (-0.25 ; 0.25) | 0.996 |
|  | HC and TLE | -0.09 | (-0.33 ; 0.15) | 0.695 |
|  | TLE and GGE | 0.09 | (-0.2 ; 0.39) | 0.735 |
|  | LTLE and RTLE | 0.41 | (0.02 ; 0.79) | 0.188 |
| Ambiguous | HC and GGE | 0.11 | (-0.17 ; 0.39) | 0.695 |
|  | HC and TLE | -0.03 | (-0.3 ; 0.23) | 0.863 |
|  | TLE and GGE | 0.14 | (-0.19 ; 0.48) | 0.695 |
|  | LTLE and RTLE | 0.33 | (-0.14 ; 0.81) | 0.407 |
| Intentional | HC and GGE | -0.04 | (-0.41 ; 0.33) | 0.863 |
|  | HC and TLE | -0.16 | (-0.52 ; 0.20) | 0.695 |
|  | TLE and GGE | 0.11 | (-0.33 ; 0.56) | 0.750 |
|  | LTLE and RTLE | 0.45 | (0.13 ; 1.02) | 0.348 |
| Accidental | HC and GGE | -0.07 | (-0.27 ; 0.14) | 0.726 |
|  | HC and TLE | -0.09 | (-0.31 ; 0.14) | 0.695 |
|  | TLE and GGE | 0.02 | (-0.22 ; 0.25) | 0.906 |
|  | LTLE and RTLE | 0.44 | (0.12 ; 0.77) | 0.080 |

FDR = false discovery rate

### Ambiguous Intentions Hostility Questionnaire (AIHQ)

We used the AIHQ to measure social cognitive biases in PWE. Researchers developed AIHQ to measure cognitive biases in people with schizophrenia (Combs et al., 2007). Since then, AIHQ has been used to examine other patient populations. For example, Decombe et al. used the test to assess social cognitive biases in individuals with Parkinson’s disease (Decombe et al., 2022), and Neumann et al. used it in individuals after traumatic brain injury (Neumann et al., 2020).

AIHQ consists of 15 short vignettes of social situations, differing in intentionality: five are intentional, five are accidental, and five are ambiguous. Participants always complete the questionnaire in the presence of an administrator, who supervises the completeness of their answers as needed. After each situation, the administrator presents the participant with five questions: two require a verbal response, which the administrator later rates, and three are accompanied by Likert scales. Three different administrators examined the patients: VP (supervising administrator), EG, and PK. Each administrator examined the participants alone and later discussed the ratings of verbal-response questions with the supervising administrator to minimise inter-rater variability. Administrators were not blinded due to practical limitations, as testing took place during routine clinical visits. All administrators had access to the example answers during the rating. Altogether, the questionnaire yields three scores: Aggression bias (AB), Hostility bias (HB), and Blame score (BS). Each of these scores can be further divided based on the intentionality of the situations presented: ambiguous, accidental, or intentional. Hostility bias and Aggression bias scores are derived from verbal responses using example answers as a reference. Scoring ranges from 1 to 5. They reflect the first (What do you think the real reason was why they did what they did?) and the last question (What would you do about it?). Blame score is an average of the Likert scale scores from the second, third and fourth questions (Did X do that on purpose?, How angry would this make you feel?, How much would you blame X?). The second question’s Likert scale score (Did X do that on purpose?) ranges from 1 to 6; all other scores range from 1 to 5. The higher the score, the greater the bias.

### Statistical analysis

We used Welch’s t-test to compare PWE and HC in AIHQ. For the relationship between the AIHQ score, epilepsy group, age, and education, we used a linear model, assuming different variances of scores across epilepsy groups and a common variance for age (or education). We further corrected for multiple comparisons using the Benjamini-Hochberg method to control the False Discovery Rate (FDR) at a 5% level (Benjamini and Hochberg, 2018). FDR closely aligns with our inferential goal of identifying which aspects the investigated groups differ in. Methods that control the family-wise error rate (e.g., the Bonferroni correction) are better suited for testing whether the groups differ at all. We set the threshold for statistical significance at a p-value < 0.05. All analyses were performed using R Version 4.3.3.

## 3. RESULTS

### Populations

The final number of HC participants included in the analysis was 43 (16 male, 27 female, mean age of 33.0 ± 10.8). We examined 53 PWE from the Motol Epilepsy Centre: 26 with GGE (5 male, 21 female; mean age 36.9 ± 12.4) and 27 with TLE (16 male, 11 female; mean age 41.8 ± 14.0). See Table 1a. The HC group was on average younger than the TLE group (difference: -8.8, 95% CI: -15.0 to -2.42, p = 0.007); other age comparisons were not statistically significant (HC vs GGE difference -3.8, 95% CI -9.7 to 2.1, p = 0.19, GGE vs TLE difference -4.9, 95% CI -12.2 to 2.3, p = 0.18).

Out of 26 GGE participants, 24 were diagnosed as IGE, four were categorised as generalised tonic-clonic seizures only syndrome, 13 as juvenile absence epilepsy, and seven as juvenile myoclonic epilepsy. We decided to enrol two participants with epilepsy with eyelid myoclonia, even though we are aware that, according to the new ILAE definition (Hirsch et al., 2022), they do not belong to the IGE group, but together with the IGE group, correspond to an overlapping term, genetic generalised epilepsies (GGE).

Of the 27 subjects with TLE, 23 had lesional mesial TLE (resulting from hippocampal sclerosis, malformation of cortical development, cavernoma, or cerebral contusion), and four had non-lesional TLE. Regarding laterality, 11 had right-sided, and 16 had left-sided TLE. For further, more detailed clinical characteristics of the GGE and TLE groups, see the supplementary data.

### Sociodemographic characteristics

Male and female representation differed across groups, with the TLE group having the highest proportion of male participants (HC: 37% male, GGE: 19% male, and TLE: 59% male). In the HC group, the difference in HB scores between male and female participants was not statistically significant, and the effect size was small to moderate (difference: 0.11, 95% CI: -0.05 to 0.26, p = 0.16, Hedge’s g = 0,47, 95% CI: -0.16 to 1.11). Similar results were obtained for the GGE group (difference: 0.13, 95% CI: -0.12 to 0.39, p = 0.26, Hedge’s g = 0.47, 95% CI:-0.54 to 0.39) as well as the TLE group (difference: 0.09, 95% CI: -0.14 to 0.32, p = 0.42, Hedge’s g = 0.31, 95% CI:-0.48 to 0.32).

HC and both PWE groups differed in age and education, with HC younger than both PWE groups but having higher education levels than both (see Table 1a). In a sensitivity analysis using a linear model (assuming different variances of scores within epilepsy groups), and using education, age and sex as additional predictors, we see a similar result for comparison between GGE and HC (difference: 0.25, 95% CI: 0.12 – 0.39, p < 0.001, Hedges’s g = 0.91, 95% CI: 0.43 - 1.4) as well as between TLE and HC (difference: 0.19, 95% CI: 0.05 – 0.32, p = 0.006, Hedge’s g = 0.68, 95% CI: 0.2 - 1.15) while the level of education, age and sex had smaller impact (all 95% CI within -0.03 – 0.19, all p > 0.07), thus not affecting the AIHQ scores substantially. In other words, these findings suggest that age, sex or education were not reliably associated with the AIHQ score; epilepsy diagnosis was.

### AIHQ results

#### Hostility bias (HB)

The GGE and TLE groups scored, on average, higher in HB than the HC group. The estimated difference (ED) for HC vs GGE was 0.25 (95 % CI 0.12;0.38, p < 0.05, Hedges’s g = -0.99, 95 % CI -1.51;-0.47), and for HC vs TLE 0.18 (95% CI 0.05;0.32, p = 0.059, Hedge’s g = -0.73 95% CI -1.23;-0.23). Even though the highest ED was evident in ambiguous situations for both groups specifically for HC vs GGE 0.35 (95% CI 0.12;0.58 p=0.059 Hedge’s g = -0.81 95% CI -1.32; -0.3) and for HC vs TLE 0.28 (95% CI 0.06;0.5 p=0.091 Hedge’s g = -0.64 95% CI -1.14 -0.15), these differences did not reach conventional levels of statistical significance based on p-values. Similarly, for intentional situations for the GGE group, where ED = 0.34 (95% CI 0.1;0.58, p = 0.059, Hedge’s g = -0.71, 95% CI -1.2; -0.2). However, the effect sizes were moderate to large, and the confidence intervals did not include zero, suggesting a potentially meaningful group difference. We did not find statistically significant differences between the GGE and TLE groups (all 95% CIs within -0.19 to 0.4; all p > 0.3). Additionally, an exploratory analysis was conducted to examine TLE subgroups by lesion laterality (left-sided temporal lobe epilepsy [LTLE] vs right-sided temporal lobe epilepsy [RTLE]). No statistically significant differences were observed between LTLE and RTLE in HB scores, and all corresponding effect size confidence intervals included zero.

#### Aggression bias (AB)

The GGE and TLE groups scored, on average, slightly higher in aggression bias than the HC group. The differences were minimal, not statistically significant and most evident in ambiguous situations (GGE vs HC ED 0.26, 95% CI 0.07;0.46, p = 0.072 Hedge’s g = -0.70 95% CI -1.21; -0.2, TLE vs HC ED 0.20, 95% CI 0.01;0.4, p = 0.176, Hedge’s g = -0.54 95% CI -1.04; -.05). No statistically significant differences were observed across subscores (all 95% CIs within -0.29 – 0.5, all p > .07), suggesting that any effects are likely small. Similarly, no differences were observed between the GGE and TLE groups (all 95% CIs within -0.41–0.47, all p > 0.16). Additionally, we conducted an exploratory analysis to examine TLE subgroups by lesion laterality. No statistically significant differences were observed between LTLE and RTLE in AB scores, and all corresponding effect size confidence intervals included zero.

#### Blame score

No statistically significant differences in blame scores were observed between groups (all 95% CI within -0.51 – 0.55, all p > 0.69), suggesting that any effects are likely small. The GGE group showed slightly higher mean scores than the HC and TLE groups, with the largest differences observed in ambiguous situations; however, these differences were not statistically significant. Additionally, an exploratory analysis was conducted to examine TLE subgroups based on lesion laterality. No statistically significant differences were observed between LTLE and RTLE in blame scores. For a comprehensive overview of effect size measures, see the supplement.

#### Epilepsy-related variables and AIHQ scores

For this analysis, we did not divide PWE into GGE and TLE subgroups because the subgroups were small. Although the GGE and TLE groups were similar in age (GGE 36.9 ± 12.4, TLE 41.8 ± 14.0), their disease duration differed (GGE 22.2 ± 13.0, TLE 11.7 ± 4.5). This difference is not surprising, as the onset of GGE is typically in early adolescence. We found no correlations between disease duration and AIHQ scores.

The PWE were divided into three groups: those without medication (2 participants), those with 1 antiseizure medication (28 participants), and those with> 1 antiseizure medication (23 participants). For the statistical model, PWE without medication were excluded. We observed no difference in PWE AIHQ scores related to their medication. Additionally, we compared PWE treated with either levetiracetam, brivaracetam, or both to PWE treated with a different medication. We did not find large differences between groups (all 95% CIs were within -0.23 to 0.39, and all *p-values* were greater than *0*.*34*).

Similarly, we found no strong effect of seizures on performance in the AIHQ. However, we observed a positive correlation between subjectively evaluated depressive symptoms in PWE, measured by NDDI-E and Blame scores, in the AIHQ (95% CI for change in BS per 10-point increase in NDDI-E: 0.28–1.1, *p = 0*.*016*).

## 4. DISCUSSION

According to Fiske and Taylor, social cognition incorporates “how people make sense of other people and themselves to coordinate with their social world” (Fiske and Taylor, 2013). To understand the world around us, we must understand the causes of others’ behaviour. This knowledge fosters a sense of security in a predictable environment, promoting prosocial behaviour and a sense of belonging to a social group. Systematically erring in these judgements could result in social maladaptation. Our study demonstrates higher hostile attributional bias scores in GGE than in HC, with larger average estimated differences in ambiguous situations. This finding is consistent with other patient populations where attributional bias has been examined, such as schizophrenia (Combs, 2007) or Parkinson’s disease (Decombe, 2022), as ambiguous situations are those where situational cues are lacking. Ambivalent situations reflect one’s beliefs and values in how others are perceived, creating space for social cognitive biases (Výrost and Slaměník, 1997).

The GGE and TLE groups showed higher hostility bias scores than the HC group, but only the difference between the GGE and HC groups was statistically significant. Hostility bias is a tendency to interpret others’ behaviour as hostile, even when social cues are absent. Hostility bias has been connected to impulsivity, negative emotional experiences, and problematic or even potentially aggressive behaviour (Chen et al., 2012). Impulsivity is characterised by quick responses to stimuli without evaluating the consequences of behaviours (Chen et al., 2012). People with GGE, mainly juvenile myoclonic epilepsy (JME) (Shakeshaft et al., 2021), have increased impulsivity and deficits in executive functions. Moschetta et al. specifically linked these deficits to social maladjustment (Moschetta and Valente, 2013). Lee et al. found a higher prevalence of impulsivity in people with IGE and frontal lobe epilepsy than in people with TLE (Lee et al., 2022), further supporting the association between frontal lobe dysfunctions and impulsivity. Studies suggest that people with TLE exhibit varying degrees of social cognition deficits, primarily in ToM and ER (Bora and Meletti, 2016). Research that compared people with IGE and TLE reported higher levels of social cognition impairment for TLE groups (Bora and Meletti, 2016; Gomez-Ibañez et al., 2014; Morou et al., 2018; Realmuto et al., 2015a; Tallarita et al., 2023), highlighting the importance of the lesions in the temporal lobes in areas necessary for ToM and ER (Realmuto et al., 2015b).

According to the fMRI study by Kestemont et al., brain areas involved in attributing causes to oneself, others, or situations include the right temporal pole, bilateral temporoparietal junction, precuneus, and adjacent posterior cingulate cortex, as well as the left posterior superior temporal sulcus, left angular gyrus, and left middle temporal gyrus. The task in their study was to attribute the cause of a situation based on a one-sentence claim, such as “Someone lies to you” (Kestemont et al., 2015b). However, the TLE group in this study consists of people with predominantly (23/27) lesional mesial temporal lobe epilepsy with MRI-verified lesions in the hippocampus or amygdala, sparing the neocortical lateral regions. Ma et al. used a different paradigm to map the causal attribution network. The instruction was to infer others’ personality traits, and the dorsal medial prefrontal cortex was also recruited (Ma et al., 2012), suggesting the importance of both frontal and temporal regions in causal attributions. Although nonlesional brain MRI is traditionally considered crucial for diagnosing GGE, recent research has revealed several structural and functional network abnormalities in this patient group. These include volume reductions in the whole brain, the thalamus, the putamen, the caudate, the pallidum, and the supplementary motor area. Additionally, reductions in grey matter volume have been observed in both hemispheres, the thalamus, and the insula, as well as a decrease in surface area in the caudal anterior cingulate cortex. Structural abnormalities probably extend beyond these regions, involving attention and other cognitive functions (Loughman et al., 2014; Nuyts et al., 2017).

### Demographic characteristics and the AIHQ results

Studies on hostile attributional bias are inconsistent in terms of the effects of either male or female sex on tendencies to overinterpret situations as hostile events. One reason for inconsistencies is the underrepresentation of female participants in studies focused on hostility and aggression (Tuente et al., 2019). Chen et al. reported that males tend to externalise their anger, leading to aggression; however, when faced with an ambiguous situation, females were more likely to respond angrily (Chen et al., 2012). Populations in this study varied in female-to-male ratios, with the highest male proportion in the TLE group and the lowest in the GGE group. If male sex were seen as a major contributor to hostile attributional bias, higher scores would be expected in the TLE group. However, the highest hostility bias scores were observed in the GGE group despite its lower male proportion. This pattern is consistent with the statistical model used in this study, in which sex was not a significant predictor of HB scores. These findings suggest that sex differences are unlikely to account for the increased hostility bias observed in the GGE group.

Evidence explaining the relationship between education level and hostile attributional bias is scarce. Data from developmental psychological studies suggest that verbal competence and language development, which are typically associated with education, are important predictors of future hostile attributional bias. At the same time, authors underline the importance of cognitive flexibility, emotional regulation, and Theory of Mind abilities for social competence (Choe et al., 2013). Indirect evidence from studies examining hostile attributional bias in highly educated populations (e.g., university students) suggests that higher education is not a preventive factor against a hostile attributional style (Bailey and Ostrov, 2008), indicating that other factors play an important role. In our study, the HC group had higher levels of education than the TLE or GGE groups, with GGE the least educated, achieving the highest HB scores. Despite these group differences, the sensitivity analysis found no statistically significant effect of education on HB. This finding could support the interpretation that the effect of education is likely present, but probably indirect and small, mediated by other variables such as language or Theory of Mind skills.

Hostile attributional bias is common in young children, possibly due to limited capacity for emotion regulation and an underdeveloped ability to differentiate between others’ mental states and their own (Theory of Mind) (Choe et al., 2013). From childhood to early adolescence, as social competence grows, hostility bias appears to be attenuated and more context-dependent. However, it does not follow a linear pattern (Castro et al., 2002). In this study, the TLE group was, on average, older than the GGE and HC groups. Yet the GGE group obtained higher HB scores. Sensitivity analysis did not find a statistically significant effect of age on HB scores. Our results could support the interpretation that the relationship between age and HB is likely nonlinear, consistent with findings from studies of youth.

### Epilepsy-related variables and the AIHQ results

Out of the 53 PWE, only 15 experienced seizures 30 days before the examination. Due to heterogeneity across groups, we created a dichotomous variable indicating whether individuals had experienced seizures within 30 days before AIHQ test administration. Even though the higher frequency of seizures is associated with social maladaptation (Moschetta and Valente, 2013), we did not find any correlation between seizure occurrence and attributional bias results.

Regarding anti-seizure medication, authors often hypothesise about the potentially harmful effects of medication on social cognition in PWE. Stewart et al. found lower social competence in children with GGE who were taking lamotrigine and ethosuximide, and reduced performance in the faux pas task in children with GGE who were using valproate. (Moschetta and Valente, 2013; Stewart et al., 2018) Levetiracetam, brivaracetam, perampanel, and topiramate have been linked to behavioural adverse events, mainly hostility. (Steinhoff et al., 2021) We did not confirm these results, even when comparing PWE using levetiracetam or brivaracetam to PWE using a different medication.

We measured depressive symptoms with the Czech version of NDDI-E, where, according to the meta-analysis by Kim et al. (Kim et al., 2019), the cut-off score possibly indicating major depressive disorder is 13 points. NDDI-E is a screening tool and does not replace the clinical diagnosis of major depressive disorder. Depressive symptoms are connected with hostility and anger, mainly towards oneself, usually represented by exaggerated self-guilt and self-criticism (Moreno et al., 1994). Literature describes the so-called “self-blaming bias” - an attributional style in which people tend to blame themselves for the adverse events in their lives, regardless of the external circumstances (Kestemont et al., 2015). Of the 53 patients, 17 (7 with TLE and 10 with GGE) reached or exceeded this cut-off score. Depressive symptoms are highly prevalent in PWE (Mula et al., 2021). Kanner et al. even proposed a bidirectional relationship between depression and epilepsy based on a possible common neuropathological mechanism (Kanner, 2011). In this study, contrary to self-blaming bias, depressive symptoms were positively correlated with the blame score in the AIHQ in PWE. The blame score comprises Likert scale scores of three questions mapping the overt hostility of an individual (Did X do that on purpose?, How angry would this make you feel?, How much would you blame X?). Our results surprisingly suggest the tendency of PWE with depressive symptoms to blame external circumstances for their misfortunes rather than blaming themselves.

One possible explanation could be rooted in learned helplessness theory, which posits that individuals who repeatedly experience adverse events beyond their control (e.g., seizures in PWE) tend to expect mostly adverse outcomes in everyday situations (Vargas and Arnett, 2013). Hermann et al. investigated the relationship between attributional style and depression in people with poorly controlled TLE. Their results support a connection between depression and maladaptive pessimistic attributional style originating from learned helplessness with emphasis on internalised hostility (Hermann et al., 1996). However, their method for investigating attributional style, using the Optimism-Pessimism scale developed by Colligan et al., differed from that employed in this study. It was based on the currently outdated version of the Minnesota Multiphasic Personality Inventory. Participants were provided with statements and instructed to answer dichotomously (yes/no). In contrast, in the AIHQ, questions are more specific and may prompt more hostile responses (e.g., “How angry would this make you feel?”).

Another explanation is that the possibly exaggerated NDDI-E scores may result from the measurement technique. Subjective scales like NDDI-E could falsely aggravate the depressive symptoms in PWE, reflecting more on their frustration with suffering from a chronic health condition than on the depression itself. Less severe forms of depression have been associated with hostility towards the environment. (Moreno et al., 1994)

### Limitations and prospects

This study has several limitations. Our study samples were relatively small and heterogeneous with respect to GGE syndromes, TLE aetiology, seizure frequency, and antiseizure medication. The HC group did not match the epilepsy groups in age, education and sex distribution. Due to underpowered sample sizes, we pooled the TLE and GGE groups for analysis of epilepsy-related variables and AIHQ scores, potentially introducing bias. We did not provide an objective measurement of depression, such as the Montgomery-Asberg Depression Rating Scale. We did not collect psychosocial variables that could influence or result from a maladaptive attributional style in PWE, such as perceived stigma, employment status, marital status, or parental style. Parental overprotectiveness and anxiety are associated with lower quality of life in children with epilepsy (Jones and Reilly, 2016) and can influence social engagement in PWE (Szemere and Jokeit, 2015). The stigma of epilepsy is strongly associated with social isolation (Yücesoy and Canbolat, 2025), quality of life, perceived helplessness, and, according to Hermann et al., one of the predictors for psychopathology in PWE (Hermann et al., 1990; Jacoby et al., 2005). People with epilepsy are at higher risk of suffering from psychiatric conditions than the general population (Mula et al., 2021). Specifically, JME is associated with a higher frequency of personality disorders (Filho and Yacubian, 2013; Panda et al., 2023). Borderline personality disorder has been associated with both impulsivity and attributional bias (Schulze et al., 2024; Ursulet et al., 2023). Future studies on attributional styles in people with epilepsy could also examine personality disorders and introduce psychiatric history, family history and psychiatric evaluation into the analysis.

To better establish relationships with other social cognition frameworks, such as ToM, ER, and attributional bias, participants could also be tested using established measures, e.g., the Faux Pas test or the Pictures of Facial Affect by Ekman and Friesen. Jeon et al. used AIHQ, Brüne’s Theory of Mind Picture Stories task, and neurocognitive tests in a healthy population. Their results suggest that attributional style is primarily influenced by ToM skills, rather than cognitive functions such as working memory or attention (Jeon et al., 2012). Furthermore, attributional bias testing could help monitor the outcomes of social cognition therapeutic interventions in the future. Even though there are no gold standard therapeutic options for social cognitive impairments in PWE, there have been therapeutic attempts in different patient populations. Kurtz et al. discussed the efficacy of psychosocial interventions in individuals with schizophrenia, where Social Cognition and Interaction Training [61], Social Cognitive Skills Training [62], or the Metacognitive and Social Cognitive Skills Program [63] have shown small to moderate effect sizes on aggression, blame, and hostility, respectively. All cited authors used AIHQ as the outcome measure (Kurtz et al., 2016).

## 5. CONCLUSION

In summary, our results suggest that people with GGE may be more prone to hostile attributional bias than HC individuals. Due to considerable limitations, these results should be interpreted as preliminary, and further research on this topic is warranted, as its findings can have a significant impact on the real life of PWE. Notably, aberrant attributional styles have been successfully therapeutically influenced in various patient populations.

## Data Availability

All data produced in the present study are available upon reasonable request to the authors.

## 7. ACKNOWLEDGEMENTS

Author Viktoria Pytelova was supported by the Ministry of Health, Czech Republic, for the conceptual development of the research organisation, Motol University Hospital, Prague, Czech Republic, under grant 00064203 - JUNIOR GRANT.

Author Viktoria Pytelova was supported by Charles University, project GAUK No 169124, and the Epilepsy Research Centre in Prague (EpiReC).

Author Jana Amlerova was supported by IPE No. 699012 and the National Institute for Neurological Research No. LX22NPO5107, funded by the EU’s Next Gen EU.

Author Michaela Kalinova was supported by the ERDF-Project Brain dynamics, No. CZ.02.01.01/00/22_008/0004643

The authors thank Dr Jana Zarubova and Dr Petra Kloudova for their help in identifying participants enrolled in this research.

## 8. DECLARATION OF GENERATIVE AI AND AI-ASSISTED TECHNOLOGIES IN THE WRITING PROCESS

During the preparation of this work, the authors utilised Grammarly, Inc., to enhance readability. After using this tool, the authors reviewed and edited the content as necessary and took full responsibility for the publication’s content.

## REFERENCES

Amlerova, J., Cavanna, A.E., Bradac, O., Javurkova, A., Raudenska, J., Marusic, P., 2014. Emotion recognition and social cognition in temporal lobe epilepsy and the effect of epilepsy surgery. Epilepsy Behav 36, 86–89. 10.1016/j.yebeh.2014.05.001

Association, A.P., 2013. Diagnostic and Statistical Manual of Mental Disorders, DSM-5. 10.1176/appi.books.9780890425596

Bailey, C.A., Ostrov, J.M., 2008. Differentiating Forms and Functions of Aggression in Emerging Adults: Associations with Hostile Attribution Biases and Normative Beliefs. J. Youth Adolesc. 37, 713–722. 10.1007/s10964-007-9211-5

Benjamini, Y., Hochberg, Y., 2018. Controlling the False Discovery Rate: A Practical and Powerful Approach to Multiple Testing. J. R. Stat. Soc. Ser. B: Stat. Methodol. 57, 289–300. 10.1111/j.2517-6161.1995.tb02031.x

Berg, A.T., Vickrey, B.G., Testa, F.M., Levy, S.R., Shinnar, S., DiMario, F., 2007. Behavior and social competency in idiopathic and cryptogenic childhood epilepsy. Dev. Med. Child Neurol. 49, 487–492. 10.1111/j.1469-8749.2007.00487.x

Bora, E., Meletti, S., 2016. Social cognition in temporal lobe epilepsy: A systematic review and meta-analysis. Epilepsy Behav. 60, 50–57. 10.1016/j.yebeh.2016.04.024

Broicher, S.D., Kuchukhidze, G., Grunwald, T., Krämer, G., Kurthen, M., Jokeit, H., 2012. “Tell me how do I feel” – Emotion recognition and theory of mind in symptomatic mesial temporal lobe epilepsy. Neuropsychologia 50, 118–128. 10.1016/j.neuropsychologia.2011.11.005

Buck, B., Iwanski, C., Healey, K.M., Green, M.F., Horan, W.P., Kern, R.S., Lee, J., Marder, S.R., Reise, S.P., Penn, D.L., 2017. Improving measurement of attributional style in schizophrenia; A psychometric evaluation of the Ambiguous Intentions Hostility Questionnaire (AIHQ). J. Psychiatr. Res. 89, 48–54. 10.1016/j.jpsychires.2017.01.004

Castro, B.O.D., Veerman, J.W., Koops, W., Bosch, J.D., Monshouwer, H.J., 2002. Hostile Attribution of Intent and Aggressive Behavior: A Meta‐Analysis. Child Dev. 73, 916–934. 10.1111/1467-8624.00447

Chen, P., Coccaro, E.F., Jacobson, K.C., 2012. Hostile Attributional Bias, Negative Emotional Responding, and Aggression in Adults: Moderating Effects of Gender and Impulsivity. Aggress. Behav. 38, 47–63. 10.1002/ab.21407

Choe, D.E., Lane, J.D., Grabell, A.S., Olson, S.L., 2013. Developmental Precursors of Young School-Age Children’s Hostile Attribution Bias. Dev. Psychol. 49, 2245–2256. 10.1037/a0032293

Combs, D.R., Penn, D.L., Wicher, M., Waldheter, E., 2007. The Ambiguous Intentions Hostility Questionnaire (AIHQ): A new measure for evaluating hostile social-cognitive biases in paranoia. Cogn Neuropsychiatry 12, 128–143. 10.1080/13546800600787854

Decombe, L., Henry, A., Decombe, R., Tir, M., Maindreville, A.D.de, Hairabedian, L.G., Kaladjian, A., Raucher-Chéné, D., 2022a. “Accidental, really?” Attributional bias in patients with Parkinson’s disease. Parkinsonism Relat D 95, 18–22. 10.1016/j.parkreldis.2021.12.013

Decombe, L., Henry, A., Decombe, R., Tir, M., Maindreville, A.D. de, Hairabedian, L.G., Kaladjian, A., Raucher-Chéné, D., 2022b. “Accidental, really?” Attributional bias in patients with Parkinson’s disease. Parkinsonism Relat D 95, 18–22. 10.1016/j.parkreldis.2021.12.013

Farrant, A., Morris, R.G., Russell, T., Elwes, R., Akanuma, N., Alarcón, G., Koutroumanidis, M., 2005. Social cognition in frontal lobe epilepsy. Epilepsy Behav 7, 506–516. 10.1016/j.yebeh.2005.07.018

Filho, G.M. de A., Yacubian, E.M.T., 2013. Juvenile myoclonic epilepsy: Psychiatric comorbidity and impact on outcome. Epilepsy Behav. 28, S74–S80. 10.1016/j.yebeh.2013.03.026

Fiske, S.T., Taylor, S.E., 2013. Social Cognition: From Brains to Culture. SAGE.

Gehlert, S., 1994. Perceptions of Control in Adults with Epilepsy. Epilepsia 35, 81–88. 10.1111/j.1528-1157.1994.tb02915.x

Giovagnoli, A.R., Parente, A., Ciuffini, R., Tallarita, G.M., Turner, K., Maialetti, A., Marrelli, A.M., Pucci, B., AgainstEpilepsy, the S.G. on C.N., Neuropsychology –. Adult Section of the Italian League, 2021. Diversified social cognition in temporal lobe epilepsy. Acta Neurol Scand 143, 396–406. 10.1111/ane.13386

Gomez-Ibañez, A., Urrestarazu, E., Viteri, C., 2014. Recognition of facial emotions and identity in patients with mesial temporal lobe and idiopathic generalized epilepsy: An eye-tracking study. Seizure 23, 892–898. 10.1016/j.seizure.2014.08.012

Hao, L., Yang, J., Wang, Y., Zhang, S., Xie, P., Luo, Q., Ren, G., Qiu, J., 2015. Neural correlates of causal attribution in negative events of depressed patients: Evidence from an fMRI study. Clin Neurophysiol 126, 1331–1337. 10.1016/j.clinph.2014.10.146

Hermann, B.P., Trenerry, M.R., Colligan, R.C., Consortium, T.B.E.S., 1996. Learned Helplessness, Attributional Style, and Depression in Epilepsy. Epilepsia 37, 680–686. 10.1111/j.1528-1157.1996.tb00633.x

Hermann, B.P., Whitman, S., Wyler, A.R., Anton, M.T., Vanderzwagg, R., 1990. Psychosocial Predictors of Psychopathology in Epilepsy. Br. J. Psychiatry 156, 98–105. 10.1192/bjp.156.1.98

Hirsch, E., French, J., Scheffer, I.E., Bogacz, A., Alsaadi, T., Sperling, M.R., Abdulla, F., Zuberi, S.M., Trinka, E., Specchio, N., Somerville, E., Samia, P., Riney, K., Nabbout, R., Jain, S., Wilmshurst, J.M., Auvin, S., Wiebe, S., Perucca, E., Moshé, S.L., Tinuper, P., Wirrell, E.C., Adikaibe, D.B., Baradi, R.A., Andrade, D., Bast, T., Beydoun, A., Bien, C., Caraballo, R., Coan, A.C., Connolly, M., Dunne, J., Haut, S., Jansen, F., Jobst, B., Kalviainen, R., Kakooza, A., Kato, M., Knupp, K., Kochen, S., Lagae, L., Mayor, L.C., Okujava, N., Radakishnan, K., Roulet‐Perez, E., Rios, L., Sadleir, L., Juan‐Orta, D.S., Serratosa, J., Shellhaas, R., Tsai, M., Udani, V., Zhang, H.Y., Zhou, D., 2022. ILAE definition of the Idiopathic Generalized Epilepsy Syndromes: Position statement by the ILAE Task Force on Nosology and Definitions. Epilepsia 63, 1475–1499. 10.1111/epi.17236

Jacoby, A., Snape, D., Baker, G.A., 2005. Epilepsy and social identity: the stigma of a chronic neurological disorder. Lancet Neurol. 4, 171–178. 10.1016/s1474-4422(05)01014-8

Jeon, I.H., Kim, K.R., Kim, H.H., Park, J.Y., Lee, M., Jo, H.H., Koo, S.J., Jeong, Y.J., Song, Y.Y., Kang, J.I., Lee, S.Y., Lee, E., An, S.K., 2012. Attributional Style in Healthy Persons: Its Association with “Theory of Mind” Skills. Psychiatry Investig. 10, 34–40. 10.4306/pi.2013.10.1.34

Jokeit, H., 2010. 12. Social cognition in patients with temporal lobe epilepsies. Epilepsy Behav. 17, 582–583. 10.1016/j.yebeh.2010.01.037

Jones, C., Reilly, C., 2016. Parental anxiety in childhood epilepsy: A systematic review. Epilepsia 57, 529–537. 10.1111/epi.13326

Kanner, A.M., 2022. Suicidality in Patients With Epilepsy: Why Should Neurologists Care? Frontiers Integr Neurosci 16, 898547. 10.3389/fnint.2022.898547

Kanner, A.M., 2011. Depression and epilepsy: A bidirectional relation? Epilepsia 52, 21–27. 10.1111/j.1528-1167.2010.02907.x

Kestemont, J., Ma, N., Baetens, K., Clément, N., Overwalle, F.V., Vandekerckhove, M., 2015a. Neural correlates of attributing causes to the self, another person and the situation. Soc Cogn Affect Neur 10, 114–121. 10.1093/scan/nsu030

Kestemont, J., Ma, N., Baetens, K., Clément, N., Overwalle, F.V., Vandekerckhove, M., 2015b. Neural correlates of attributing causes to the self, another person and the situation. Soc Cogn Affect Neur 10, 114–121. 10.1093/scan/nsu030

Kim, D.-H., Kim, Y.-S., Yang, T.-W., Kwon, O.-Y., 2019. Optimal cutoff score of the Neurological Disorders Depression Inventory for Epilepsy (NDDI-E) for detecting major depressive disorder: A meta-analysis. Epilepsy Behav. 92, 61–70. 10.1016/j.yebeh.2018.12.006

Krendl, A.C., Betzel, R.F., 2022. Social cognitive network neuroscience. Soc. Cogn. Affect. Neurosci. 17, 510–529. 10.1093/scan/nsac020

Kurtz, M.M., Gagen, E., Rocha, N.B.F., Machado, S., Penn, D.L., 2016. Comprehensive treatments for social cognitive deficits in schizophrenia: A critical review and effect-size analysis of controlled studies. Clin. Psychol. Rev. 43, 80–89. 10.1016/j.cpr.2015.09.003

Lee, S.-A., Yang, H., Im, K., Choi, E.J., Jeon, J.-Y., Han, S.-H., Kim, H.-W., Lee, G.-H., Ryu, H.U., 2022. Comparisons of impulsivity among patients with different subtypes of epilepsy. Epilepsy Res. 186, 106997. 10.1016/j.eplepsyres.2022.106997

Loughman, A., Bowden, S.C., D’Souza, W., 2014. Cognitive functioning in idiopathic generalised epilepsies: A systematic review and meta-analysis. Neurosci. Biobehav. Rev. 43, 20–34. 10.1016/j.neubiorev.2014.02.012

Mirabel, H., Guinet, V., Voltzenlogel, V., Pradier, S., Hennion, S., 2020. Social cognition in epilepsy: State of the art and perspectives. Rev Neurol 176, 468–479. 10.1016/j.neurol.2020.02.010

Moreno, J.K., Selby, M.J., Fuhriman, A., Laver, G.D., 1994. Hostility in Depression. Psychol. Rep. 75, 1391–1401. 10.2466/pr0.1994.75.3.1391

Morou, N., Papaliagkas, V., Markouli, E., Karagianni, M., Nazlidou, E., Spilioti, M., Afrantou, T., Kimiskidis, V.K., Foroglou, N., Kosmidis, M.H., 2018. Theory of Mind impairment in focal versus generalized epilepsy. Epilepsy Behav 88, 244–250. 10.1016/j.yebeh.2018.09.026

Moschetta, S., Valente, K.D., 2013. Impulsivity and seizure frequency, but not cognitive deficits, impact social adjustment in patients with juvenile myoclonic epilepsy. Epilepsia 54, 866–870. 10.1111/epi.12116

Mula, M., Kanner, A.M., Jetté, N., Sander, J.W., 2021. Psychiatric Comorbidities in People With Epilepsy. Neurol. Clin. Pr. 11, e112–e120. 10.1212/cpj.0000000000000874

Neumann, D., Sander, A.M., Perkins, S.M., Bhamidipalli, S.S., Witwer, N., Combs, D., Hammond, F.M., 2020. Assessing Negative Attributions After Brain Injury With the Ambiguous Intentions Hostility Questionnaire. J. Head Trauma Rehabilitation 35, E450– E457. 10.1097/htr.0000000000000581

Nuyts, S., D’Souza, W., Bowden, S.C., Vogrin, S.J., 2017. Structural brain abnormalities in genetic generalized epilepsies: A systematic review and meta‐analysis. Epilepsia 58, 2025– 2037. 10.1111/epi.13928

Ogurcakova, V., Kajsova, M., Marusic, P., Amlerova, J., 2024. Social cognition in Idiopathic generalised epilepsies. Behav. Brain Res. 469, 115044. 10.1016/j.bbr.2024.115044

Panda, P.K., Ramachandran, A., Tomar, A., Elwadhi, A., Kumar, V., Sharawat, I.K., 2023. Prevalence, nature, and severity of the psychiatric comorbidities and their impact on quality of life in adolescents with Juvenile myoclonic epilepsy. Epilepsy Behav. 142, 109216. 10.1016/j.yebeh.2023.109216

Parsons, N., Bowden, S.C., Vogrin, S., D’Souza, W.J., 2020. Default mode network dysfunction in idiopathic generalised epilepsy. Epilepsy Res. 159, 106254. 10.1016/j.eplepsyres.2019.106254

Realmuto, S., Zummo, L., Cerami, C., Agrò, L., Dodich, A., Canessa, N., Zizzo, A., Fierro, B., Daniele, O., 2015a. Social cognition dysfunctions in patients with epilepsy: Evidence from patients with temporal lobe and idiopathic generalized epilepsies. Epilepsy Behav 47, 98– 103. 10.1016/j.yebeh.2015.04.048

Realmuto, S., Zummo, L., Cerami, C., Agrò, L., Dodich, A., Canessa, N., Zizzo, A., Fierro, B., Daniele, O., 2015b. Social cognition dysfunctions in patients with epilepsy: Evidence from patients with temporal lobe and idiopathic generalized epilepsies. Epilepsy Behav 47, 98– 103. 10.1016/j.yebeh.2015.04.048

Schachter, S.C., 2006. Quality of life for patients with epilepsy is determined by more than seizure control: the role of psychosocial factors. Expert Rev. Neurother. 6, 111–118. 10.1586/14737175.6.1.111

Schulze, A., Rommelfanger, B., Schendel, E., Schott, H., Lerchl, A., Vonderlin, R., Lis, S., 2024. Attributional style in Borderline personality disorder is associated with self-esteem and loneliness. Borderline Pers. Disord. Emot. Dysregulation 11, 19. 10.1186/s40479-024-00263-2

Shakeshaft, A., Panjwani, N., McDowall, R., Crudgington, H., Ceballos, J.P., Andrade, D.M., Beier, C.P., Fong, C.Y., Gesche, J., Greenberg, D.A., Hamandi, K., Koht, J., Lim, K.S., Orsini, A., Rees, M.I., Rubboli, G., Selmer, K.K., Smith, A.B., Striano, P., Syvertsen, M., Talvik, I., Thomas, R.H., Zarubova, J., Richardson, M.P., Strug, L.J., Pal, D.K., Consortium, the B., 2021. Trait impulsivity in Juvenile Myoclonic Epilepsy. Ann. Clin. Transl. Neurol. 8, 138–152. 10.1002/acn3.51255

Smith, H.L., Summers, B.J., Dillon, K.H., Macatee, R.J., Cougle, J.R., 2016. Hostile interpretation bias in depression. J. Affect. Disord. 203, 9–13. 10.1016/j.jad.2016.05.070

Steiger, B.K., Jokeit, H., 2017. Why epilepsy challenges social life. Seizure 44, 194–198. 10.1016/j.seizure.2016.09.008

Steinhoff, B.J., Klein, P., Klitgaard, H., Laloyaux, C., Moseley, B.D., Ricchetti-Masterson, K., Rosenow, F., Sirven, J.I., Smith, B., Stern, J.M., Toledo, M., Zipfel, P.A., Villanueva, V., 2021. Behavioral adverse events with brivaracetam, levetiracetam, perampanel, and topiramate: A systematic review. Epilepsy Behav. 118, 107939. 10.1016/j.yebeh.2021.107939

Stewart, E., Catroppa, C., Gill, D., Webster, R., Lawson, J., Mandalis, A., Sabaz, M., Barton, B., Lah, S., 2018. Theory of Mind and social competence in children and adolescents with genetic generalised epilepsy (GGE): Relationships to epilepsy severity and anti-epileptic drugs. Seizure 60, 96–104. 10.1016/j.seizure.2018.06.015

Stewart, E., Catroppa, C., Gonzalez, L., Gill, D., Webster, R., Lawson, J., Sabaz, M., Mandalis, A., Barton, B., McLean, S., Lah, S., 2019. Facial emotion perception and social competence in children (8 to 16 years old) with genetic generalized epilepsy and temporal lobe epilepsy. Epilepsy Behav 100, 106301. 10.1016/j.yebeh.2019.04.054

Szemere, E., Jokeit, H., 2015. Quality of life is social – Towards an improvement of social abilities in patients with epilepsy. Seizure 26, 12–21. 10.1016/j.seizure.2014.12.008

Tallarita, G.M., Ciuffini, R., Turner, K., Pucci, B., Parente, A., Giovagnoli, A.R., 2023. Social Cognition in Idiopathic Generalized Epilepsies and Nonlesional Temporal Lobe Epilepsy. Acta Neurol. Scand. 2023, 1–11. 10.1155/2023/9450272

Tuente, S.K., Bogaerts, S., Veling, W., 2019. Hostile attribution bias and aggression in adults - a systematic review. Aggress Violent Beh 46, 66–81. 10.1016/j.avb.2019.01.009

Ursulet, A., Repentigny, É. de Quansah, J.E., Bessette, M., Gagnon, J., 2023. An International Collection of Multidisciplinary Approaches to Violence and Aggression. 10.5772/intechopen.105367

Vargas, G.A., Arnett, P.A., 2013. Attributional Style and Depression in Multiple Sclerosis. Int. J. MS Care 15, 81–89. 10.7224/1537-2073.2012-021

Výrost, J., Slaměník, I., 1997. Sociální psychologie. ISV - nakladatelství, Praha.

Yogarajah, M., Mula, M., 2019. Social cognition, psychiatric comorbidities, and quality of life in adults with epilepsy. Epilepsy Behav 100, 106321. 10.1016/j.yebeh.2019.05.017

Yücesoy, M., Canbolat, O., 2025. The relationship between disease concealment, stigmatization, and self-management in adult patients with epilepsy in Turkey. Seizure: Eur. J. Epilepsy. 10.1016/j.seizure.2025.08.021

Zanão, T.A., Lopes, T.M., Campos, B.M. de, Yasuda, C.L., Cendes, F., 2021. Patterns of default mode network in temporal lobe epilepsy with and without hippocampal sclerosis. Epilepsy Behav. 121, 106523. 10.1016/j.yebeh.2019.106523

Zhang, T., Chen, L., Wang, Y., Zhang, M., Wang, L., Xu, X., Xiao, G., Chen, J., Shen, Y., Zhou, N., 2018. Impaired theory of mind in Chinese children and adolescents with idiopathic generalized epilepsy: Association with behavioral manifestations of executive dysfunction. Epilepsy Behav 79, 205–212. 10.1016/j.yebeh.2017.12.006

Ziaei, M., Arnold, C., Thompson, K., Reutens, D., 2022a. Social Cognition in Temporal and Frontal Lobe Epilepsy: Systematic review, Meta-analysis, and Clinical Recommendations. Medrxiv 2021.04.28.21255765. 10.1101/2021.04.28.21255765

Ziaei, M., Arnold, C., Thompson, K., Reutens, D.C., 2022b. Social Cognition in Temporal and Frontal Lobe Epilepsy: Systematic Review, Meta-analysis, and Clinical Recommendations. J Int Neuropsych Soc 1–25. 10.1017/s1355617722000066

